# Exploring Community Perspectives on Barriers and Facilitators to Childhood Immunization among First Nations Peoples in Northern Saskatchewan: A Qualitative Sharing Circle Study

**DOI:** 10.1101/2025.10.08.25337612

**Authors:** Nnamdi Ndubuka, Emmanuel Dankwah, Priscilla Dankwah, Shirley Woods, Grace Akinjobi, Carrie Gardipy-Mckenzie, Justina Ndubuka, Adeshola Abati, Amanda Froehlich-Chow, Geoffrey Maina, Ibrahim Khan, Gary Groot

## Abstract

Childhood vaccination is vital for disease prevention intervention, yet some Northern Saskatchewan First Nations communities have immunization coverage below the 95% target. We aimed to explore community perspectives on the barriers and facilitators of childhood immunization uptake in First Nations communities in Northern Saskatchewan. We used a qualitative design informed by Indigenous methodologies and community-based participatory approaches to engage three First Nations communities and conducted sharing circles to gather rich, culturally grounded perspectives on childhood immunization. We purposefully recruited parents, caregivers, an Elder, and a community leader. We audio-recorded and transcribed one sharing circle (5 participants) discussion, then analyzed data thematically with Indigenous research principles, ensuring community collaboration and ethical stewardship throughout the study. The results were validated through a community feedback session. Participants described several structural and social factors that limited access to immunization, including transportation barriers, limited clinic hours, geographic isolation, and reduced services during the COVID-19 pandemic. Caregiver mental health issues, substance use, and unstable home environments were identified as barriers to immunization uptake. Language barriers, lack of health literacy, and historical trauma also shaped vaccine hesitancy. Key enablers identified included establishining trusting relationships with local healthcare providers, use of mobile clinics, peer support, tailored communication strategies, community events that incorporated immunization programs rooted in cultural practices, and involvement of Elders and local leadership in planning and delivery. Our findings show that vaccine uptake in Northern Saskatchewan First Nations communities reflects a complex set of contextual factors, shaped by structural conditions and historical experiences. We recommend policy and practice approaches that support culturally safe, community-led immunization programming. Strengthening trust, expanding flexible service models, and integrating First Nations knowledge systems into public health planning can help improve childhood vaccination coverage and contribute to health equity in northern and remote settings.

## Introduction

Childhood immunization remains one of the most impactful public health strategies for preventing infectious diseases and safeguarding population health. Routine vaccination programs have drastically reduced the burden of diseases such as measles, pertussis, and polio. According to the World Health Organization (1), immunization averts an estimated 2 to 3 million deaths globally each year. Vaccines not only provide direct protection to children but also support herd immunity, reducing transmission and protecting individuals who cannot be immunized due to medical conditions (2,3).

Despite the well-documented benefits of vaccination, coverage remains uneven across different populations in Canada. Childhood vaccines are publicly funded and universally available through the National Immunization Program, but significant disparities persist particularly among Indigenous populations, leaving them more vulnerable to vaccine-preventable diseases (4)(4). In 2021, only 82.5% of Indigenous children were fully vaccinated for measles, mumps, and rubella (MMR) by age two, compared to 91.6% of all Canadian children. For diphtheria, tetanus, and pertussis (DTaP), 67.4% of Indigenous two-year-olds were fully vaccinated, compared to 77.1% of their non-Indigenous peers. Similarly, coverage for the Haemophilus influenzae type b (Hib) vaccine among Indigenous children was 65.3%, significantly lower than the national rate of 75.3% (5,6). These lower coverage rates are reflected in disproportionately higher rates of vaccine-preventable diseases among Indigenous children and adults compared to non-Indigenous Canadians (4,7,8).

These disparities are rooted in broader structural and historical inequities. Geographic isolation, socioeconomic disadvantage, limited health infrastructure, and jurisdictional complexities between federal and provincial healthcare responsibilities all contribute to inconsistent access to timely and culturally appropriate immunization services (9). Moreover, legacies of colonization, including the residential school system and forced assimilation policies, have fostered mistrust in mainstream healthcare systems among many Indigenous communities (10,11).

First Nations communities in Saskatchewan exemplify the ongoing challenges in achieving equitable childhood immunization coverage. Vaccination rates in these communities are notably lower than both the provincial averages and the national target of 95%. As of June 2022, the percentage of children in Saskatchewan who had received the recommended number of doses by their second birthday was 73.4% for pertussis, 73.3% for measles, and 85.6% for meningococcal serogroup C (12). In comparison, among First Nations children living in First Nations communities, coverage was significantly lower, 43.2% for pertussis, 43% for measles, and 74.5% for meningococcal serogroup C (12). Many of these communities are served by the Northern Inter-Tribal Health Authority (NITHA) and are located in remote and rural areas of the province. These locations face numerous barriers to consistent immunization delivery, including challenging weather conditions, limited transportation infrastructure, and insufficient healthcare services (13–15).

Despite persistent challenges, some First Nations communities in Northern Saskatchewan have demonstrated success through community-led approaches that prioritize cultural safety and self-determination (15). These initiatives have effectively increased immunization uptake by embedding Indigenous languages, traditions, and leadership into health promotion. For example, some communities have produced vaccine education materials in Cree and Dene, hosted clinics in culturally significant spaces such as band halls or community centers, and opened events with prayers or smudging ceremonies. Elders and community champions play an active role in their outreach, offering trusted guidance and personal stories to build vaccine confidence. The sustained partnerships between Band Councils and the health authority have further supported the alignment of public health efforts with community values and priorities (15). These approaches align with broader principles of Indigenous self-determination, relational accountability, and the importance of centering Indigenous knowledge in health policy and practice (16). Since 2015, NITHA has formally recognized these efforts through its *Childhood Immunization Coverage Awards*, conferred upon communities achieving immunization rates of 90% or higher (15).

Although these are indeed examples of positive development, significant knowledge gaps remain. Surveillance systems often capture quantitative immunization data, but they do not adequately reflect the contextual and experiential factors that influence caregiver decision-making. Understanding why immunization rates remain low in some communities requires listening to the voices of those most directly affected parents, caregivers, Elders, and community leaders. Community-engaged and qualitative research approaches are essential for uncovering the nuanced realities, cultural values, and systemic barriers that shape immunization behaviours.

Our study was therefore designed to explore community perspectives on childhood immunization uptake within selected Northern Saskatchewan First Nations. We aimed to pinpoint community-defined barriers and enablers that shape vaccine acceptance and vaccine series completion for children aged 0–2 years. This age group was prioritized because it represents a critical window for routine immunizations that protect against several serious, vaccine-preventable diseases. Delays or gaps in immunization during this period can significantly increase vulnerability to illness and contribute to broader public health disparities, particularly in communities already experiencing systemic barriers to healthcare access. We sought not only to generate evidence rooted in the lived experiences of community members, but also to guide the development and delivery of immunization programs that are culturally informed and tailored to the specific social and contextual realities of First Nations communities. Our study answered the question: What community-identified barriers and enablers influence caregivers’ decision to vaccinate First Nations children aged 0-2 years in Northern Saskatchewan.

## Materials and Methods

### Study Design

We adopted a qualitative descriptive methodology underpinned by thematic analysis to explore community perspectives on childhood immunization. Guided by Indigenous research methodologies, our study was informed by Two-Eyed Seeing, a framework that brings together the strengths of Indigenous and Western ways of knowing (17). Anchored in a community-based research framework, we prioritized First Nations voices and partnership which guided all phases of the research process (18). To create a culturally responsive approach, we utilized Indigenous sharing circle techniques, a recognized method for facilitating open, respectful, and collective dialogue in Indigenous contexts (19). We employed sharing circles within a broader focus group discussion structure to promote storytelling and co-learning among participants in a safe and culturally grounded environment.

Consistent with the First Nations principles of OCAP®—Ownership, Control, Access, and Possession, we ensured that participating communities retained authority over their information and its use in the research (20). This included co-developing the research design, co-facilitating data collection processes, and validating findings collaboratively. Acknowledging the challenges of ensuring anonymity in qualitative research within close-knit communities, we applied strict confidentiality measures and collaborated with community leaders to uphold both participant privacy and community control. Relational accountability, an Indigenous research principle that emphasizes the importance of relationships, respect, and responsibility, was embedded throughout the project to ensure that all knowledge shared was treated ethically and reciprocally (21).

### Study Setting

We conducted this study in Northern Saskatchewan, Canada, within the jurisdiction of NITHA. NITHA is a First Nations-driven health organization that provides public health and primary care support to 33 on-reserve First Nations communities across northern Saskatchewan. NITHA plays a pivotal role in delivering immunization programs and other population health services in partnership with community-based health providers. These communities are geographically dispersed, often remote or semi-isolated, and many face challenges related to health care access, service delivery infrastructure, and social determinants of health (13–15,22).

The region is characterized by significant health disparities, a challenge commonly faced by many Indigenous communities across Canada (10). Immunization coverage in many NITHA communities remains below provincial and national targets, contributing to elevated risks for vaccine-preventable diseases among First Nations children (23,24).

We invited three First Nations communities to participate in the study based on their relatively low immunization rates as indicated by the NITHA immunization surveillance data and their willingness to support research activities. Community leadership provided formal letters of support affirming their collaboration in the project and the importance of addressing childhood vaccination challenges. The setting was thus purposefully chosen to reflect both the community’s need for enhanced immunization uptake and their willingness to co-lead research rooted in Indigenous knowledge systems.

### Study Participants

Participants in this study were members of First Nations communities, with a focus on parents and caregivers responsible for childhood immunization decisions. We used purposeful sampling to recruit individuals with relevant lived experience and insight into immunization-related behaviors and challenges (25). Community health workers played a leading role in the recruitment process, disseminating invitations via word-of-mouth and email. Participant receruitment occurred between July 1, 2024 and September 10, 2024. The recruitment approach aligns with Indigenous research methodologies that emphasize relationship-building, trust, and culturally safe engagement (26).

We assembled a final participant group of five individuals: one Elder, three caregivers (each representing a participating community), and one community leader (Table 1). This composition captured traditional knowledge, day-to-day caregiving experience, and community-governance insight. The inclusion of an Elder in our study facilitated the integration of cultural and historical contexts into our interpretation of immunization experiences and barriers, thereby honoring Indigenous protocols that emphasize intergenerational knowledge-sharing (27).

Although the participant group may be small, this aligns with Indigenous qualitative methodologies, which emphasize relationality, trust, and cultural safety. These principles support meaningful, community-informed dialogue (19).

**Table 1.** Participant Characteristics (n = 5)

| Characteristics | Category | Number of participants (n) | Percentage (%) |
| --- | --- | --- | --- |
| Gender | Female | 4 | 80% |
|  | Male | 1 | 20% |
| Age | Adult (≥ 18 years) | 5 | 100% |

### Data Collection

We gathered stories (qualitative data) through a culturally grounded sharing circle held, at Wanuskewin Heritage Park, an Indigenous heritage site of deep cultural and historical significance located in Northern Saskatchewan. We used sharing circle approach, which is widely recognized as a traditional Indigenous method of group dialogue to foster open, respectful, and inclusive conversations among participants (19). A First Nations Elder opened the gathering with prayer, establishing a respectful, safe space for dialogue in keeping with Indigenous protocols (26). After a concise overview of local childhood-immunization trends and challenges, we invited participants for a one-hour sharing circle discussion, creating a safe space for community members to share their stories and insights on barriers and enablers of vaccine uptake in the participating First Nations communities.

We obtained informed written consent from every participant, ensuring that each person understood the study’s purpose, procedures, and voluntary nature, including the right to withdraw at any time without consequence. Recognizing that the discussion topics might evoke emotional discomfort, we put culturally appropriate supports in place: an Indigenous Elder remained on site throughout the session, and we informed participants about the Hope for Wellness Helpline for immediate mental-health or crisis assistance.

Research assistants who self-identified as First Nations and were trained in qualitative methods and Indigenous research ethics facilitated the sharing circle. With participants’ consent, we audio-recorded the session. Two team members then transcribed the recordings verbatim to preserve participants’ exact words, and First Nations assistants translated any Indigenous-language segments into English to maintain meaning and context. We also recorded non-verbal cues and key discussion points in field notes and on flip charts. The complete discussion guide is provided in the supporting information (S1 Table).

We upheld confidentiality at every stage of data collection. Before the session, we reminded participants to respect one another’s privacy and keep the conversation confidential. Afterward, we locked hard-copy documents in a double-locked cabinet and stored digital files on a password-protected computer, then transferred them to a secure University of Saskatchewan OneDrive. Only authorized team members could access these materials, ensuring participant privacy and meeting data-governance standards.

### Data Analysis and Reporting

We conducted a thematic analysis informed by phenomenological principles to explore participants’ lived experiences and perceptions regarding childhood immunization within their communities (29). This approach allowed us to identify patterns of meaning across the dataset while preserving the depth and context of individual narratives. Members of the research team independently reviewed and coded the verbatim transcripts of the sharing circle discussions. We used both manual techniques and NVivo 14 qualitative analysis software to support systematic coding and data organization.

The analytical process involved iterative reading of the transcripts, followed by the identification and grouping of significant statements into initial codes. These codes were then clustered into overarching themes and subthemes that captured the complexity and nuance of participants’ experiences. The themes were refined through team discussions to ensure coherence, relevance, and alignment with the study objectives. To protect the anonymity of participants, we reported all findings in aggregate form without attributing specific quotes to identifiable individuals.

To enhance the trustworthiness and credibility of the results, we implemented a member-checking process through a community-based knowledge translation (KT) event held in Prince Albert, Saskatchewan. During this event, we presented preliminary findings to participants and key community stakeholders. This session provided an opportunity for attendees to reflect on the interpretations, clarify meanings, and offer feedback. We integrated this input into the final analysis to ensure that the findings authentically represented the participants’ voices and community perspectives. This validation process also reinforced our commitment to relational accountability and co-creation of knowledge with Indigenous communities. Through this approach, we aimed to produce findings that are not only methodologically rigorous but also socially and culturally meaningful to the communities involved.

### Ethical Considerations

We conducted this study in accordance with established ethical guidelines for research involving human participants and Indigenous communities. We respected Indigenous cultural protocols at each stage of engagement. The leadership of the Northern Inter-Tribal Health Authority approved the project, enabling us to engage in initiatives that will significantly benefit the communities. Our research project followed the principles of the Tri-Council Policy Statement (TCPS-2), “Chapter 9: Research Involving the First Nations, Inuit, and Metis Peoples of Canada” (28) and the First Nations principles of Ownership, Control, Access, and Power (OCAP) (20) to ensure that the research was used to derive positive impacts for communities while preserving traditional ways of knowing and governance. We obtained written consent from all participants. The University of Saskatchewan’s Behavioural Research Ethics Board approved the study (ID# Beh-REB4894).

## Results

### Barriers to Childhood Immunization

#### Access and Logistics

Participants identified transportation as a critical barrier due to the remote locations of homes and lack of available transport to health centers. One participant explained the challenge by stating, *“For sure…the transportation and, you know, a lot of our young moms…don’t have vehicles and live far away from the clinic.”* The geographical distance between homes and health facilities, combined with limited or unavailable public transport, further complicated access to immunization services.

Additionally, participants noted that the scheduling of immunization clinics typically limited to morning hours, did not align with the daily routines of many families, particularly younger parents. Morning appointments were often described as difficult to attend. One participant noted, “*A lot of our young people, they don’t get up till mid-afternoon*,” while another remarked, “*It’s like they’re up all night, sleep all day*.” The study participants attributed the irregular sleep pattern to the influence of the local casino culture. Several participants suggested that late-night gambling contributed to disrupted household routines and inconsistent sleep schedules. As one caregiver explained, “*I think gambling probably would be [a factor], because there are casinos that are open pretty much 24/7*.” These findings show a clear disconnect between the operating hours of immunization clinics and the lived realities of community members, potentially limiting access to timely vaccination services.

#### Knowledge and Communication Gaps

Participants expressed concern about the limited information provided to parents regarding childhood immunizations. Many felt that education was either missing or insufficient at key points, such as during hospital discharge or early postpartum visits. One caregiver reflected, *“Lack of education from when they had the baby at the hospital or the nurse….”* Others noted a lack of follow-up and continued guidance, with one participant stating, *“Lack of ongoing education to parents… a better awareness of clinics… you don’t see that happening anymore.”* This gap in consistent communication contributed to confusion, uncertainty, and missed appointments.

Language emerged as a significant barrier, with participants emphasizing the difficulties posed by limited English proficiency and complex medical terminology. One participant explained vividly, *“Language barrier…If you don’t speak good English, you’re not gonna understand. But if I say it in Cree, our language is so powerful…you will understand me better.”* Another emphasized the use of simpler language and visual aids, stating, *“They use big words…if that doesn’t work, the last card is draw pictures.”* Participants highlighted the critical need for culturally and linguistically appropriate health communication to improve understanding and engagement.

#### Household and Lifestyle Challenges

Participants frequently discussed the interconnected challenges of substance use, unstable home environments, and associated feelings of shame and social isolation as significant barriers to childhood immunization.

Substance use disorders emerged as a primary factor. Participants explained that widespread substance abuse, especially alcoholism and crystal meth, created difficult family environments and often disrupted healthcare-seeking behaviors. One participant explained explicitly: *“Alcoholism, drug abuse… mostly crystal meth… not only the parents of their child, but it could be the grandparents, like the home they live in.”*

Chaotic Home Environments were also noted, often resulting from or exacerbated by substance use. Participants described these environments as unstable, unpredictable, and disruptive to routine care, including immunization visits.

Further, participants highlighted how feelings of Shame and Social Isolation tied to home life and substance use significantly impacted clinic attendance. Families were described as reluctant to expose their home circumstances or lifestyles to outsiders, including healthcare providers. One participant shared: *“I’m thinking because of their home lifestyle…that shame of the home lifestyle, or they don’t wanna leave their little safe haven.”* This sentiment was echoed by another, who emphasized the emotional toll of judgment: *“A lot of people aren’t comfortable…having someone [health workers] at the end of the night.”* The deeply intertwined nature of shame and addiction was clearly articulated by another participant: *“It’s a lot of shame within the addiction… that’s where I see the problem is in our community.”*

#### Social and Cultural Influences

Family and peer beliefs played a profound role in shaping immunization decisions. As one participant described, *“If your mom and dad had never had the vaccination, then it’s not important to them,”* further explaining the role of generational beliefs: *“I’ve never heard my mom saying, you gotta go get your vaccination.”* These attitudes were seen to spread within families and peer networks, reinforcing negative or indifferent attitudes toward immunization.

Participants also emphasized the deep impact of cultural beliefs, historical trauma, and the influential role of Elders on immunization uptake. Experiences from residential schools left lasting impacts, causing deep mistrust toward health interventions. As one participant explained: *“For residential school survivors…they had no choice…it traumatized them so much that they didn’t want to put their children through what they went through.”* Additionally, religious and cultural beliefs compounded skepticism toward vaccines.

Further complicating the issue, some Elders, highly respected within the community, expressed uncertainty or hesitation toward vaccines. A participant explicitly noted, *“A lot of Elders do not believe in vaccination…although elders are very highly respected in the communities.”* Another participant attributed this skepticism partly to lack of information, stating, *“The lack of education with elders not knowing what this is about. What’s this gonna do? But then again, what are the long-term effects of the immunizations?”* These intertwined factors significantly shaped immunization decisions across generations, suggesting that addressing Elders’ perspectives is vital to improving vaccination uptake.

#### Psychological and Competing Demands

Mental health challenges emerged as significant obstacles to accessing immunizations. One participant described the complexity, noting, *“If a person has mental issues…it’s really hard to talk to the person, even to deal with their cancer because they’re so into their mental disability.”* Others affirmed the impact of parental mental health conditions like anxiety and depression, highlighting, *“Because of people’s anxiety and depression, they themselves can’t get their child out.”*

Daily survival priorities frequently took precedence over preventive healthcare. As one participant pointed out *“Their priorities [are] not trying to get their baby to get a needle. Their priorities [are] what are they gonna eat for supper?”* These competing demands often pushed immunization to the background, as immediate needs like food and shelter commanded urgent attention.

Participants also spoke to the challenges faced by young parents, many of whom lack parenting knowledge and support networks. One participant succinctly described this as a *“lack of parenting,”* while another illustrated the daily struggles young families encounter: *“In a lot of young families, young moms, they’re really struggling…their priorities, not trying to get their baby to get a needle. Their priorities, what are they gonna eat for supper?”* This reflects broader issues of resource scarcity and competing survival priorities that overshadow preventive health care.

#### Health System Shortfalls

Participants emphasized a marked decline in proactive clinic outreach and follow-up efforts. One participant noted, *“A better awareness from the clinic…maybe have more home visits. You don’t see that happening anymore.”* Others suggested practical reasons for declining home visits, such as personal discomfort with visits due to shame or judgment: *“When it comes to home visits, a lot of people don’t…want them to see [their] house is messy.”*

The COVID-19 pandemic significantly disrupted immunization services and community engagement. One participant noted the severe disruption, stating, *“COVID really dampened a lot of things. We all know that.”* Another explained further, *“We lost all that with COVID because everybody was so scared of the needle… there’s just no discussion at this time.”* Participants shared that the pandemic profoundly affected attitudes towards healthcare visits, leading some to avoid clinics entirely.

### Enablers of Childhood Immunization

#### Accessible Service Design

Participants emphasized the value of phone call reminders as simple yet effective strategies to increase immunization attendance. The reminders served as a practical and reliable means of communication for busy families. As one participant explained, *“They let you know. Like, they’ll phone, and they’ll be like, hey, so-and-so needs a 3-month immunization. Can you do it on this day?”*

The deployment of mobile clinics, which brought immunization services directly into communities, was praised for addressing geographic and transportation-related barriers. Participants appreciated the convenience and practicality of mobile health services. One participant described the model positively, stating, *“Mobile van/clinic…we bring nurses or doctors, whoever wants to come…and it’s a closed area for private people who wanna talk privately…we’re bringing the clinic to the people.”*

Integrating immunization services with community-based social events, such as baby showers, fostered welcoming and non-clinical environments that encouraged participation among parents and caregivers. These gatherings facilitated informal health conversations within a culturally resonant context. As one participant suggested: *“I would have, like, even a shower for all the young moms…and we’ll eat together. Because food is such a comfort zone for all of us. And when you’re eating, you’re talking, … and we start talking about vaccines.”*

#### Effective Communication Channels

Participants identified social media platforms particularly Facebook and community radio as essential channels for disseminating immunization-related information and updates. These mediums were commonly relied upon to stay informed about local health services and community events. As one participant observed, *“Advertising on Facebook…all young people use Facebook. Social media.”* Another participant emphasized the continued relevance of radio-based outreach: *“We advertise on the radio. That’s how we tell our young people.”*

In addition to formal channels, informal peer-to-peer communication within the community emerged as a critical means of promoting vaccine awareness and uptake. referenced the role of interpersonal dialogue in shaping vaccine decisions, particularly among parents. One participant described the impact of such interactions, noting the influence of a *“sharing circle…listening to [another mother] in a sharing circle”* as influential in vaccine decision-making.

#### Trust and Social Support

rusted community members, family, and friends played a pivotal role in encouraging immunization acceptance. Participants emphasized the power of role modeling, noting that visible, positive examples within their networks influenced personal decisions. As one participant stated, *“Set a good example. Set your own leaders.”* Another participant reflected on how familial influence shaped their choice: *“I followed my cousin—she said it[immunization] was important.”*

Strong, trust-based relationships with healthcare providers also significantly shaped vaccination behaviors. Participants expressed greater willingness to vaccinate when providers were familiar, consistent, and embedded in the fabric of community life. One participant captured this dynamic: *“The relationship…the trust…there’s one of our nurses that’s been there for years. Everybody could go through her. That’s everybody’s grandma… we grew up with her.”* In contrast, participants described lower immunization uptake in contexts where provider relationships were weak or transient.

Participants advocated for peer involvement, particularly younger parents who had successfully navigated the immunization process. While perspectives on elder involvement varied, many valued the combination of peer relatability and elder wisdom. One participant explained, *“Peer support with some elder involvement…I would rather go with that young mom that had the experience…but I would also involve the elder for support.”*

#### Motivation and Engagement

Incentives such as food, prize draws, and recreational activities effectively motivated families to attend immunization events. Participants consistently described these strategies as practical and appealing, contributing to increased participation and community enthusiasm. As one participant noted, *“Food incentives, bingo, draws…prizes would get people to come in.”* Such incentives not only encouraged attendance but also created a more positive, engaging environment around vaccination.

Active participation by clinic staff in community life further strengthened trust and encouraged ongoing engagement. When healthcare providers particularly nurses were visible and involved in local activities, community members felt more connected and confident in accessing services. One participant emphasized the value of this approach: *“Just getting involved, having those nurses in the community participating, getting to know them.”*

**Table 2.** Thematic Summary of Caregiver-Identified Barriers and Enablers to Childhood Immunization in Northern Saskatchewan First Nations.

| Barriers | Enablers |
| --- | --- |
| **Access & Logistics**<br>• Transportation challenges<br>• Morning-only clinic hours mis-aligned with household routines (casino nightlife) | **Accessible Service Design**<br>• Phone-call reminders<br>• Mobile “van” clinics<br>• Baby-shower / events for vaccination |
| **Knowledge & Communication Gaps**<br>• Limited education at hospital discharge/post-partum<br>• Lack of follow-up/outreach<br>• Language and health-literacy barriers | **Effective Communication Channels**<br>• Facebook posts & community radio<br>• Word-of-mouth & sharing-circle discussions |
| **Household & Lifestyle Challenges**<br>• Substance-use disorders (alcohol, crystal-meth)<br>• Chaotic / unstable home environments<br>• Shame-based social isolation | **Trust & Social Support**<br>• Role-modelling by peer parents<br>• Long-serving, trusted community nurses<br>• Supportive Elder involvement |
| **Social & Cultural Influences**<br>• Family / peer discouragement<br>• Historical trauma & inter-generational mistrust<br>• Elder skepticism toward vaccines | **Motivation & Engagement**<br>• Food, bingo, and prize incentives<br>• Active clinic-staff presence at community events |
| **Psychological & Competing Demands**<br>• Mental Health Issues<br>• Survival priorities (food, housing)<br>• Young/inexperienced parenting |  |
| **Health System Shortfalls**<br>• Decline in home-visit outreach<br>• Limited clinic follow-up<br>• COVID-19 Pandemic-related service disruption |  |

## Discussion

In our study, we explored the factors shaping childhood immunization uptake in Northern Saskatchewan First Nations communities, with attention to structural, social, psychological, and cultural-historical influences. Our findings build on and extend current evidence related to immunization practices in First Nations and rural Canadian settings.

Transportation challenges persist as a major barrier for families living far from health centers without reliable vehicles. These logistical hurdles, coupled with limited clinic hours that often do not align with families’ work or caregiving responsibilities, reduce opportunities for timely vaccination (30,31). Similar findings from rural settings confirm that geographic distance and inflexible service schedules contribute to missed appointments and lower immunization rates (32,33). Strategies such as mobile clinics and extended hours, including evenings and weekends, have proven successful in other contexts by bringing services closer to families and increasing accessibility (34).

The COVID-19 pandemic further limited access to care by reducing in-person interactions and increasing isolation. Global studies have documented sharp declines in routine vaccinations during the pandemic, as families avoided healthcare facilities due to fear of infection (35,36). Re-engaging families with the health system and addressing service disruptions remain essential to restoring vaccination coverage.

Participants shared several stories describing impacts of social and environmental conditions negatively affecting caregivers’ ability to maintain immunization schedules. These included substance use, gambling, unstable housing, and high levels of household stress. Previous research shows that poverty, addiction, and unstable environments create barriers to healthcare access and continuity (30,37). Feelings of shame and fear of being judged because of living conditions further discouraged some families from seeking care. This finding aligns with literature describing how stigma and discrimination continue to deter Indigenous peoples from engaging with healthcare services (16,38). Programs that offer non-judgmental and respectful services can reduce these barriers and improve service uptake.

Communication difficulties stemming from language loss and low health literacy also played a role. The decline of First Nations languages and limited access to clear, comprehensible health information can hinder understanding of immunization schedules and vaccine safety. Health promotion tools that use plain language and visual materials have shown success in similar populations (39). Incorporating First Nations languages and culturally meaningful messaging into education campaigns could improve vaccine-related communication and foster greater engagement.

Caregivers experiencing mental health challenges, including anxiety and depression, also reported difficulty attending appointments. These challenges are known to influence adherence to preventive health services, including immunizations (40). Embedding mental health screening and support in immunization programs may help address these barriers and improve participation.

Younger and less experienced parents described feeling uncertain about how to manage health responsibilities. Other research has found that adolescent and young adult parents benefit from targeted health education and mentorship to support child health and immunization (41). Tailored programs for young parents could provide essential information, guidance, and emotional support to improve vaccine adherence.

Another barrier reported by many participants was lack of trust in the health system rooted in experiences of historical trauma, including the legacy of residential schools. Some families declined immunizations based on ongoing skepticism toward Western healthcare. This mistrust reflects intergenerational experiences of coercion and marginalization within healthcare systems (11,38). These dynamics continue to shape health-related decision-making and demand approaches that recognize and respond to their origins.

Researchers have called for health services that respect Indigenous knowledge, affirm cultural identity, and support self-determination (42,43). Programs that work in partnership with Elders and community leaders can help foster trust and ensure that immunization efforts are culturally relevant and accepted (44–46). Integrating traditional healing alongside biomedical approaches has also shown promise in improving healthcare relationships and outcomes.

We identified several factors that supported vaccine uptake from the perspectives of caregivers, Elders, and community leaders. Participants consistently pointed to the importance of trusting relationships with healthcare providers. Familiarity with local staff and respectful communication encouraged participation in immunization services. This finding aligns with earlier work showing that trust, built through consistent and culturally appropriate interactions, plays a central role in immunization decisions (3,46). The principle of relational accountability, in which providers maintain meaningful and reciprocal relationships with communities, appeared important to positive immunization experiences (47).

Community-based activities such as baby showers and health-focused local events also supported vaccine access. These events provided informal settings for families to connect with health workers and created a positive environment for discussing immunization. Other research has documented similar benefits from community-led health initiatives, which strengthen local ownership and align with social norms (47,48). Offering immunization at familiar venues or in conjunction with cultural events helped normalize the process and reduce logistical challenges.

Participants said that social media, radio announcements, posters, and word of mouth kept them informed about immunization opportunities. Facebook and local radio were particularly valued in reaching families in remote areas. These findings align with public health literature advocating for multichannel communication strategies that reflect local preferences and infrastructure limitations (49–51).

Peer influence and role modeling emerged as additional enablers. Participants mentioned being influenced by family members and respected individuals who shared their own decisions to vaccinate. Prior studies show that trusted social networks and peer behaviors often guide health decision-making, especially in cultures where collective values are strong (52,53). Engaging local champions in immunization campaigns may be an effective way to promote vaccine acceptance and address hesitancy.

Other practical support included appointment reminders by phone, mobile clinics, flexible clinic hours, and incentives such as food or small prizes. These measures helped families overcome transportation and scheduling challenges, both of which are well documented in rural and Indigenous health literature (9,10,51). Mobile clinics in particular played a key role in reaching households that otherwise faced difficulty accessing services.

Taken together, our findings point to the value of health services that are community-led, flexible, and grounded in meaningful relationships. Top-down models that overlook the lived realities and cultural frameworks of First Nations families are unlikely to succeed. Instead, we support the development of immunization programs that respect Indigenous authority, center community voices, and adapt to the unique contexts of northern and remote communities (44).

### Study Strength and Limitations

One of the key strengths of our study is using a community-engaged qualitative approach, which allowed us to capture rich, in-depth insights directly from parents, caregivers, and Elders in Northern Saskatchewan First Nations communities. By engaging diverse voices, we ensured that the findings reflected a wide range of lived experiences and perspectives related to childhood immunization. The presence of an Elder at every of the engagement sessions to guide the sessions in a culturally appropriate way ensured our discussion and findings are culturally grounded and fit to be implemented in First Nations communities. We collaborated with local health leaders and community representatives throughout the study process, from design and recruitment to data interpretation. This approach enhanced the cultural relevance, credibility, and trustworthiness of our findings. Our team also included First Nations and non-First Nations researchers, which strengthened our ability to approach the data through both culturally grounded and health system lenses. We conducted interviews and focus groups in familiar, community-based settings, which fostered open dialogue and increased participant comfort. In addition, we applied a strengths-based lens, which not only identified barriers but also highlighted local enablers and community-driven solutions. This orientation allowed us to center First Nations knowledge and practices in discussions of immunization and public health programming.

Our study is not without limitations. While we captured a range of perspectives, our sample size was relatively small and may not fully reflect the diversity of experiences within and across First Nations communities. Our findings may not be generalizable to all Indigenous communities in Canada. Additionally, as with all qualitative research, the findings reflect the interpretations of the researchers and participants at the time of the study. Although we employed reflexivity and member checking to enhance rigor, some nuances may still have been missed or interpreted differently in other contexts.

Future research should include a broader range of communities and consider mixed-methods or longitudinal designs to validate and expand on these findings.

### Implications for Policy and Practice

In our study, we identified structural and social barriers that limit childhood immunization uptake in Northern Saskatchewan First Nations communities. To respond effectively, we propose that health authorities and First Nations leadership co-develop flexible immunization services such as mobile clinics, home visits, and extended hours that align with local needs and daily routines. These approaches can reduce logistical barriers and improve accessibility for families living in remote areas.

We support the integration of community-led and culturally safe practices into routine care by training healthcare providers in trauma-informed approaches and encouraging the recruitment of First Nations staff. Building respectful and consistent relationships between providers and community members remains central to increasing participation in immunization programs.

We also call for communication strategies that reflect local language use, oral traditions, and varying levels of health literacy. Using trusted community networks, radio, and social media can help share vaccine information and clinic updates more effectively.

To strengthen outcomes, we recommend linking immunization programs with mental health, parenting support, and early childhood services. These partnerships can provide holistic support for families navigating complex life circumstances. By aligning public health strategies with community-driven priorities, we contribute to improving vaccine coverage and supporting First Nations’ health sovereignty in Northern Saskatchewan.

### Conclusion

We examined the complex factors shaping childhood immunization in Northern Saskatchewan First Nations communities in our study. We found that access challenges, social conditions, historical trauma, and provider relationships all influence vaccine uptake. We also identified community-based strategies that support participation, including trusted relationships, flexible service delivery, and culturally rooted communication. We recommend that public health programs center First Nations leadership, align services with community needs, and integrate relational and culturally safe approaches to improve immunization coverage in this context.

## Data Availability

All relevant data are within the manuscript and its Supporting Information files.

## Acknowledgements

The authors gratefully acknowledge Northern Inter-Tribal Health Authority and partner community health teams for their hard work and contribution to this article. We acknowledge contributions from Shree Lamichhane, Eden Foreman, Pauline Dreaver, Barbra Crookedneck, Patricia Mckenzie, Terri Gardiner, Girija Nair and Elder Emile Highway.

## References

1. WHO. Immunization coverage. [Internet]. 2023 [cited 2025 Apr 11]. Available from: https://www.who.int/news-room/fact-sheets/detail/immunization-coverage

2. Anderson RM, May RM. Vaccination and herd immunity. Lancet. 1999;335:641–5.

3. MacDonald NE. Vaccine hesitancy: Definition, scope and determinants. Vaccine. 2015;33(34):4161–4.

4. PHAC. Vaccination coverage goals and vaccine preventable disease reduction targets by 2025. [Internet]. Ottawa. ; 2017 [cited 2025 Apr 7]. Available from: https://www.canada.ca/en/public-health/services/immunization-vaccine-priorities/national-immunization-strategy/vaccination-coverage-goals-vaccine-preventable-diseases-reduction-targets-2025.html

5. Halseth R. Vaccine Uptake among First Nations, Inuit, and Metis populations [Internet]. Prince George; 2024 [cited 2025 May 7]. Available from: https://www.nccih.ca/Publications/Lists/Publications/Attachments/10468/FS-vaccine-uptake-EN-Web.pdf

6. PHAC. Highlights From the 2021 Childhood National Immunization Coverage Survey (cNICS) [Internet]. Ottawa; 2024 [cited 2025 May 13]. Available from: https://www.canada.ca/en/public-health/services/immunization-vaccines/vaccination-coverage/2021-highlights-childhood-national-immunization-coverage-survey.html

7. Menzies RI, Singleton RJ. Vaccine preventable diseases and vaccination policy for Indigenous populations. Pediatric Clinics. 2009;56(6):1263–83.

8. Li YA, Martin I, Tsang R, Squires SG, Demczuk W, Desai S. Invasive bacterial diseases in Northern Canada, 2006-2013. Canada Communicable Disease Report. 2016;42(4):74.

9. NCCIH. Access to health services as a social determinant of First Nations, Inuit and Métis health [Internet]. National Collaborating Centre for Indigenous Health Prince George, BC, Canada; 2019 [cited 2025 Jul 18]. Available from: https://www.nccih.ca/docs/determinants/FS-AccessHealthServicesSDOH-2019-EN.pdf

10. Allan B, Smylie J. First Peoples, second class treatment: The role of racism in the health and well-being of Indigenous peoples in Canada. Wellesley Institute; 2015.

11. Greenwood M, De Leeuw S, Lindsay N. Challenges in health equity for Indigenous peoples in Canada. The Lancet. 2018;391(10131):1645–8.

12. MOH-SK. Childhood Immunization: Coverage statistics for 2- and 7-year-old children. [Internet]. 2022 [cited 2025 Jun 22]. Available from: https://www.saskatchewan.ca/residents/health/accessing-health-care-services/immunization-services/immunization-rates-in-saskatchewan

13. NITHA. Health Status Report: Social Determinants of Health Indicators [Internet]. Prince Albert, Saskatchewan; 2024 [cited 2025 Apr 29]. Available from: https://www.nitha.com/wp-content/uploads/2025/02/NITHA-HSR-2024-Chapter-3-Social-Determinants-of-Health-Indicators_20Jan25-Final.pdf

14. NITHA. Health Status Report: Organizational and Geographical Profile [Internet]. Prince Albert, Saskatchewan; 2024 [cited 2025 Apr 28]. Available from: https://www.nitha.com/wp-content/uploads/2025/02/NITHA-HSR-2024-Chapter-1-Organizational-and-Geographical-Profile_20Jan25-Final.pdf

15. NITHA. Annual Report 2023-2024 [Internet]. Prince Albert, Saskatchewan; 2024 [cited 2025 Jul 8]. Available from: https://www.nitha.com/wp-content/uploads/2024/08/NITHA-Annual-Report-2023-2024_Final_web.pdf

16. Browne AJ, Varcoe C, Lavoie J, Smye V, Wong ST, Krause M, et al. Enhancing health care equity with Indigenous populations: evidence-based strategies from an ethnographic study. BMC Health Serv Res. 2016;16(1):544.

17. Marshall M, Marshall A, Bartlett C. Two-eyed seeing in medicine. Determinants of Indigenous peoples’ health: Beyond the social. 2018;44–53.

18. Israel BA, Schulz AJ, Parker EA, Becker AB. Review of community-based research: assessing partnership approaches to improve public health. Annu Rev Public Health. 1998;19(1):173–202.

19. Lavallée LF. Practical application of an Indigenous research framework and two qualitative Indigenous research methods: Sharing circles and Anishnaabe symbol-based reflection. Int J Qual Methods. 2009;8(1):21–40.

20. FNIGC. First Nations Information Governance Centre (FNIGC). 2014 [cited 2025 May 21]. Ownership, Control, Access and Possession (OCAP®): The Path to First Nations Information Governance. Available from: https://fnigc.ca/ocap-training/

21. Wilson S. Research is ceremony: Indigenous research methods. Fernwood publishing; 2020.

22. NITHA. Northern Inter-Tribal Health Authority (NITHA). 2025 [cited 2025 May 1]. About NITHA. Available from: https://nitha.com/about/

23. NITHA. Health Status Report 2024: Immunization [Internet]. Prince Albert, Saskatchewan; 2024 [cited 2025 May 13]. Available from: https://www.nitha.com/wp-content/uploads/2025/04/NITHA-HSR-2024-Chapter-4-Immunization_15Apr25-Final.pdf.

24. NITHA. Health Status Report 2024: Communicable Disease [Internet]. Prince Albert, Saskatchewan; 2024 [cited 2025 May 15]. Available fromhttps://www.nitha.com/wp-content/uploads/2025/04/NITHA-HSR-2024-Chapter-5-Communicable-Disease_15Apr25-Final.pdf

25. Palinkas LA, Horwitz SM, Green CA, Wisdom JP, Duan N, Hoagwood K. Purposeful sampling for qualitative data collection and analysis in mixed method implementation research. Administration and policy in mental health and mental health services research. 2015;42(5):533–44.

26. Kovach M. Indigenous methodologies: Characteristics, conversations, and contexts. University of Toronto press; 2021.

27. Battiste M. Reclaiming Indigenous voice and vision. UBC press; 2011.

28. TCPS-2. Tri-Council Policy Statement: Ethical Conduct for Research Involving Humans – TCPS 2 (2022) – Chapter 2: Scope and Approach [Internet]. [cited 2025 Jun 1]. Available from: https://www.pre.ethics.gc.ca/eng/tcps2-eptc2_2022_chapter2-chapitre2.html

29. Braun V, Clarke V. Using thematic analysis in psychology. Qual Res Psychol. 2006;3(2):77– 101.

30. Reading CL, Wien F. Health inequalities and social determinants of Aboriginal peoples’ health. National Collaborating Centre for Aboriginal Health. British Columbia. 2009;

31. Okwaraji YB, Mulholland K, Schellenberg J, Andarge G, Admassu M, Edmond KM. The association between travel time to health facilities and childhood vaccine coverage in rural Ethiopia. A community based cross sectional study. BMC Public Health. 2012;12(1):476.

32. Chiem A, Olaoye F, Quinn R, Saini V. Reasons and suggestions for improving low immunization uptake among children living in low socioeconomic status communities in Northern Alberta, Canada–A qualitative study. Vaccine. 2022;40(32):4464–72.

33. MacDonald SE, Graham B, Paragg J, Foster-Boucher C, Waters N, Shea-Budgell M, et al. One child, one appointment: how institutional discourses organize the work of parents and nurses in the provision of childhood vaccination for First Nations children. Hum Vaccin Immunother. 2022;18(5):2048558.

34. LaGattuta NR, Wilson TC, Failla JA, Stoner AM, Fradua K, Brown J, et al. The effectiveness of a mobile health clinic delivering mandatory and elective middle school immunizations: A descriptive analysis. Cureus. 2023;15(9).

35. Santoli JM. Effects of the COVID-19 pandemic on routine pediatric vaccine ordering and administration—United States, 2020. MMWR Morb Mortal Wkly Rep. 2020;69.

36. Causey K, Fullman N, Sorensen RJD, Galles NC, Zheng P, Aravkin A, et al. Estimating global and regional disruptions to routine childhood vaccine coverage during the COVID-19 pandemic in 2020: a modelling study. The Lancet. 2021;398(10299):522–34.

37. Browne T, Priester MA, Clone S, Iachini A, DeHart D, Hock R. Barriers and facilitators to substance use treatment in the rural south: a qualitative study. The Journal of Rural Health. 2016;32(1):92–101.

38. Czyzewski K. Colonialism as a broader social determinant of health. The International Indigenous Policy Journal, 2 (1), 1–14. 2011.

39. Nutbeam D. The evolving concept of health literacy. Soc Sci Med. 2008;67(12):2072–8.

40. Suffel AM, Ojo-Aromokudu O, Carreira H, Mounier-Jack S, Osborn D, Warren-Gash C, et al. Exploring the impact of mental health conditions on vaccine uptake in high-income countries: a systematic review. BMC Psychiatry. 2023;23(1):15.

41. Awadh AI, Hassali MA, Al-Lela OQ, Bux SH, Elkalmi RM, Hadi H. Does an educational intervention improve parents’ knowledge about immunization? Experience from Malaysia. BMC Pediatr. 2014;14(1):254.

42. McGough S, Wynaden D, Gower S, Duggan R, Wilson R. There is no health without cultural safety: Why cultural safety matters. Contemp Nurse. 2022;58(1):33–42.

43. Nguyen HT. Patient centred care: cultural safety in indigenous health. Aust Fam Physician. 2008;37(12).

44. Henderson RI, Shea-Budgell M, Healy C, Letendre A, Bill L, Healy B, et al. First nations people’s perspectives on barriers and supports for enhancing HPV vaccination: foundations for sustainable, community-driven strategies. Gynecol Oncol. 2018;149(1):93–100.

45. Clark K, Crooks K, Jeyanathan B, Ahmed F, Kataquapit G, Sutherland C, et al. Highlighting models of Indigenous leadership and self-governance for COVID-19 vaccination programmes. AlterNative: An International Journal Of Indigenous Peoples. 2024;20(1):250–8.

46. Sanders C, Burnett K, Ray L, Ulanova M, Halperin DM, Halperin SA, et al. An exploration of the role of trust and rapport in enhancing vaccine uptake among Anishinaabe in rural northern Ontario. PLoS One. 2024;19(12):e0308876.

47. Kyoon Achan G, Eni R, Phillips-Beck W, Lavoie JG, Kinew KA, Katz A. Canada first nations strengths in Community-Based primary healthcare. Int J Environ Res Public Health. 2022;19(20):13532.

48. Nkole P, Alice F, Mutua W, Osei L, Stoljar Gold A, Yang L, et al. How a community-led understanding of access and uptake barriers and enablers informs better vaccination programs. Frontiers in Health Services. 2023;3:1260400.

49. Brusse C, Gardner K, McAullay D, Dowden M. Social media and mobile apps for health promotion in Australian Indigenous populations: scoping review. J Med Internet Res. 2014;16(12):e280.

50. Dubé E, Gagnon D, MacDonald NE. Strategies intended to address vaccine hesitancy: Review of published reviews. Vaccine. 2015;33(34):4191–203.

51. MacDonald SE, Marfo E, Sell H, Assi A, Frank-Wilson A, Atkinson K, et al. Text message reminders to improve immunization appointment attendance in Alberta, Canada: the childhood immunization reminder project pilot study. JMIR Mhealth Uhealth. 2022;10(11):e37579.

52. Brunson EK. The impact of social networks on parents’ vaccination decisions. Pediatrics. 2013;131(5):e1397–404.

53. Ghosh S, Bhattacharya S, Mukherjee S, Chakravarty S. Promote to protect: data-driven computational model of peer influence for vaccine perception. Sci Rep. 2024;14(1):306.

